# Minor Consent state policies and COVID-19 vaccination in adolescents

**DOI:** 10.64898/2026.04.10.26350608

**Authors:** Catherine Litchy, Jason Semprini

**Author notes:** **Correspondence:** Jason Semprini, 8025 Grand Ave, West Des Moines, IA 50266.

## Abstract

**Background:** Ever since the COVID-19 vaccine became available, vaccinations in adolescents lagged behind adults. Whether adolescent vaccination rates were higher in states with “Minor Consent” policies remains unknown.

**Methods:** We accessed adolescent (aged 12-17) county-level vaccine administration data from the CDC (12/2020-05/2023) Our outcomes were COVID-19 vaccination counts for 1) initial dose, 2) completed series doses, 3) booster doses. Panel Poisson regression models with state and time random effects, seasonal fixed-effects, log-population offsets, and adult vaccination rates were estimated to calculate incidence rate ratios (IRR), testing the association between residing in a state with a Minor Consent policy and COVID-19 vaccine uptake.

**Results:** Overall, for the initial dose and complete series, there was no difference in adolescent COVID-19 vaccination between states with or without Minor Consent policies. However, we found that Minor Consent policies were associated with lower COVID-19 booster doses (IRR = 0.582; 95% CI: 0.409, 0.828; p=0.0026). This association was not found in urban (IRR = 0.867; CI = 0.722, 1.043; p = 0.1295), but only in rural counties (IRR 0.541; CI 0.401, 0.730; p<0.0001).

**Conclusions:** Minor Consent policies were not associated with higher adolescent COVID-19 vaccination. Rather, we found that Minor Consent policies were associated with lower adolescent vaccination for booster doses in rural counties. Despite minimal evidence of impact, states continue to implement Minor Consent vaccination policies. Future research on the topic should investigate, not just other vaccines, but how Minor Consent policies impact parental trust in public health more broadly.

## Introduction

From the start of the COVID-19 pandemic through December 2020, the United States saw more than sixteen million cases and close to three hundred thousand deaths^1^. While new variants posed significant challenges, estimates suggest that as many as three million lives were saved within the first two years of COVID-19 vaccines being available in the US^2^. Initially authorized for limited populations, by May 2021, the FDA expanded authorization for all adolescents^3^. By Fall 2022, 71.1% of adolescents ages 12-17 had received at least one dose of a COVID-19 vaccine, and 60.8% had completed the series^4^. These rates were lower than the rates seen in adults, with the series completion rates for ages 25-49 and 50-64 being 71% and 82.7%, respectively^4^. COVID-19 vaccination rates also varied within adolescent populations^5^. Beginning early in the rollout and persisting through the pandemic, urban adolescent rates (65%) were nearly twice the rate of rural adolescents (39%)^6^.

The gap in adolescent vaccination rates prompted many to investigate parental decisions during the pandemic. Often, parent decisions for their children matched their own vaccination decisions^7^. Subsequent studies revealed the top reasons for not vaccinating adolescents included concerns about side effects, lack of trust in the COVID-19 vaccine, and lack of trust in the government^8^. General vaccine hesitancy was highest among parents in rural areas^9^.

Before the pandemic, to combat parental vaccine hesitancy and promote adolescent vaccination rates, some in public health called for policies authorizing minors to consent to receive vaccines^10^. Many states have policies that allow minors of a certain age to consent to some types of medical treatment, including vaccines^11^. A few states authorized minors to consent for COVID-19 vaccination^12^. Advocates for “Minor Consent” policies claim that adolescents develop the capacity to understand the consequences of medical decisions earlier than age eighteen and that adolescents want to be involved in medical decisions^13,14^. Research has shown that adolescents included in the COVID-19 vaccination discussion were more satisfied with their outcome, whereas parental decisions were key predictors in unvaccinated adolescents^15,16^.

However, the evidence base evaluating Minor Consent policies on vaccination outcomes has been quite limited. While these Minor Consent policies could conceptually raise vaccination rates, no study has shown such an effect for COVID-19 vaccines^14^. Only one study, to our knowledge, estimated non-causal associations between Minor Consent policies and vaccination rates; a study with misclassified state categories (i.e., Iowa authorizes minors to consent for certain vaccines but was not classified as such)^17–19^. Most of the research on this topic has, instead, involved ethics and legal perspectives^20,21^. Still, despite limited empirical evidence, many states continue debating whether to implement Minor Consent policies, either authorizing or revoking minor’s capacity to consent for vaccines^19,22,23^. Understanding how Minor Consent policies influence vaccination outcomes can help policymakers balance tradeoffs for adolescent autonomy and parental rights.

## Methods

### COVID-19 Vaccination Data

We accessed publicly available county-level COVID-19 vaccination data from the Centers for Disease Control and Prevention (CDC) COVID-19 Vaccinations in the United States, County dataset^24^. These data were compiled from jurisdictional Immunization Information Systems (IIS), aggregated vaccination records reported by multiple providers, including public health agencies, pharmacies, healthcare systems, and federal vaccination programs. These data represent a comprehensive administrative record of vaccine doses administered within a jurisdiction and are routinely reported to the CDC for surveillance and monitoring purposes. The dataset includes vaccination counts and coverage metrics across demographic groups and geographies. Our analytic sample covered the period from December 2020, corresponding to the initial rollout of COVID-19 vaccines in the United States, through May 2023, the last available date in the dataset. All vaccination measures were aggregated to the county level, by week.

### Vaccination Measures

The vaccination outcomes were weekly, county-level adolescent (ages 12–17 years) COVID-19 vaccination counts. We examined three measures of vaccination uptake: 1) Initiation: Number of adolescents receiving at least one COVID-19 vaccine dose, 2) Series completion: Number of adolescents completing the primary vaccination series, and 3) Booster uptake: Number of adolescents receiving at least one booster dose. Due to low counts, we excluded the bivalent booster dose outcomes.

All statistical models included logged adolescent population as an offset and respective COVID-19. Rural and urban status was determined by 2013 Rural-Urban Continuum Code classification (Urban = RUCC 1-3, Rural = RUCC 4-9)^25^.

### Exposure to Minor Consent Policies

State-level Minor Consent policy for COVID-19 vaccination were determined by publicly available, independent sources and then crosschecked and validated by our research team (Supplemental File 1)^26,27^. We excluded states which changed their Minor Consent policy regarding COVID-19 vaccinations during the study period (DC) and municipalities offering minors the capacity to consent for COVID-19 vaccines (Philadelphia, PA). US territories were also excluded. The states which authorized minors to consent to COVID-19 vaccinations during the study period were AR, ID, OR, RI, SC, TN, WA.

### Statistical Analysis

We estimated the association between Minor Consent policies and adolescent COVID-19 vaccination via county-week panel Poisson model. Each Poisson regression model included a logged adolescent population offset, effectively comparing rates between counties. To account for variation in COVID-19 vaccination more broadly, each model adjusted adult COVID-19 vaccination rates. To account for unobserved, time-invariant differences between states, the panel design included state and week random effects, as well as seasonal fixed-effects. Standard errors were clustered at the county level to account for within-county serial correlation over time. Results were presented as incidence rate ratios (IRRs) with corresponding confidence intervals. Subgroup analyses stratified sample by rural and urban counties. All analyses were performed in Stata v. 19.

## Results

### Summary Statistics

Table 1 presents population-adjusted mean counts of COVID-19 vaccine doses administered to adolescents aged 12–17 years at the county level between December 2020 and May 2023. The average total population-adjusted vaccinations, for all doses, were larger in states without minor consent policies than average vaccinations in states with minor consent policies.

**Table 1:** Summary Statistics – Mean Vaccination Doses.

|  |  | Initial | Complete Series | Booster |
| --- | --- | --- | --- | --- |
| Overall |  |  |  |  |
| Minor Consent |  | 4925 | 4251 | 1515 |
|  |  | [4916, 4933] | [4244, 4259] | [1511, 1520] |
| No Minor Consent |  | 5709 | 4761 | 1566 |
|  |  | [5706, 5712] | [4758, 4763] | [1565, 1568] |
| Urban |  |  |  |  |
| Minor Consent |  | 5288 | 4590 | 1716 |
|  |  | [4784, 5792] | [4091, 5088] | [1289, 2142] |
| No Minor Consent |  | 6011 | 5031 | 1684 |
|  |  | [5782, 6240] | [4841, 5220] | [1553, 1816] |
| Rural |  |  |  |  |
| Minor Consent |  | 3302 | 2742 | 622 |
|  |  | [3097, 3506] | [2562, 2922] | [540, 703] |
| No Minor Consent |  | 3670 | 2996 | 795 |
|  |  | [3435, 3905] | [2862, 3130] | [734, 855] |

Table 1 reports average total (population-adjusted) number of COVID-19 vaccine doses by county administered to adolescents aged 12-17 between December 2020 and May 2023. Data is from United States Centers for Disease Control and Prevention's Immunization Information Systems. 95% CIs are reported in brackets. All calculations derived from Poisson models and adjust for logged population offsets.

**Table 2.** Incidence Rate Ratios – Association Between State Level Minor Consent Policy and COVID-19 Vaccination.

|  |  | **Initial** | **Complete Series** | **Booster** |
| --- | --- | --- | --- | --- |
| **Overall** |  |  |  |  |
|  | Estimate | 0.858 | 0.907 | 0.582** |
|  | [95% CI] | [0.718, 1.025] | [0.706, 1.166] | [0.409, 0.828] |
|  | (p) | 0.0922 | 0.4468 | 0.0026 |
| **Urban** |  |  |  |  |
|  | Estimate | 1.003 | 0.960 | 0.867 |
|  | [95% CI] | [0.743, 1.354] | [0.447, 2.066] | [0.722, 1.043] |
|  | (p) | 0.9834 | 0.9176 | 0.1295 |
| **Rural** |  |  |  |  |
|  | Estimate | 0.871 | 0.993 | 0.541**** |
|  | [95% CI] | [0.669, 1.134] | [0.687, 1.435] | [0.401, 0.730] |
|  | (p) | 0.3063 | 0.9705 | <0.0001 |

Table 2 reports COVID-19 vaccine administration for adolescents aged 12-17 in states with and without Minor Consent policies. Data was reported by the United States Centers for Disease Control and Prevention's Immunization Information Systems between December 2020 and May 2023. Estimate represents the incidence rate ratio (IRR) of vaccine receipt for an adolescent in a state with a Minor Consent policy compared to an adolescent in a state without a Minor Consent policy. \*\* $p < 0.01$ , \*\*\*\* $p < 0.0001$

Overall, counties in states without Minor Consent laws had higher mean vaccination counts across all dose categories compared to those with Minor Consent laws. Mean initial doses were 5709 in non-Minor Consent states versus 4925 in Minor Consent states. Similarly, completed series doses averaged 4761 versus 4251, and booster doses averaged 1566 versus 1515.

These differences were more pronounced in rural counties. In rural areas, counties without Minor Consent laws had higher mean vaccination counts across all outcomes, with particularly large gaps for booster doses (795 vs. 622). In contrast, booster dose counts were similar in rural counties (non Minor Consent = 1684 vs. Minor Consent = 1716), with overlapping confidence intervals in urban counties.

Figures 1-2 show the population adjusted counts of COVID-19 vaccination doses in adolescents by exposure to minor consent policies.

**Figure 1.**
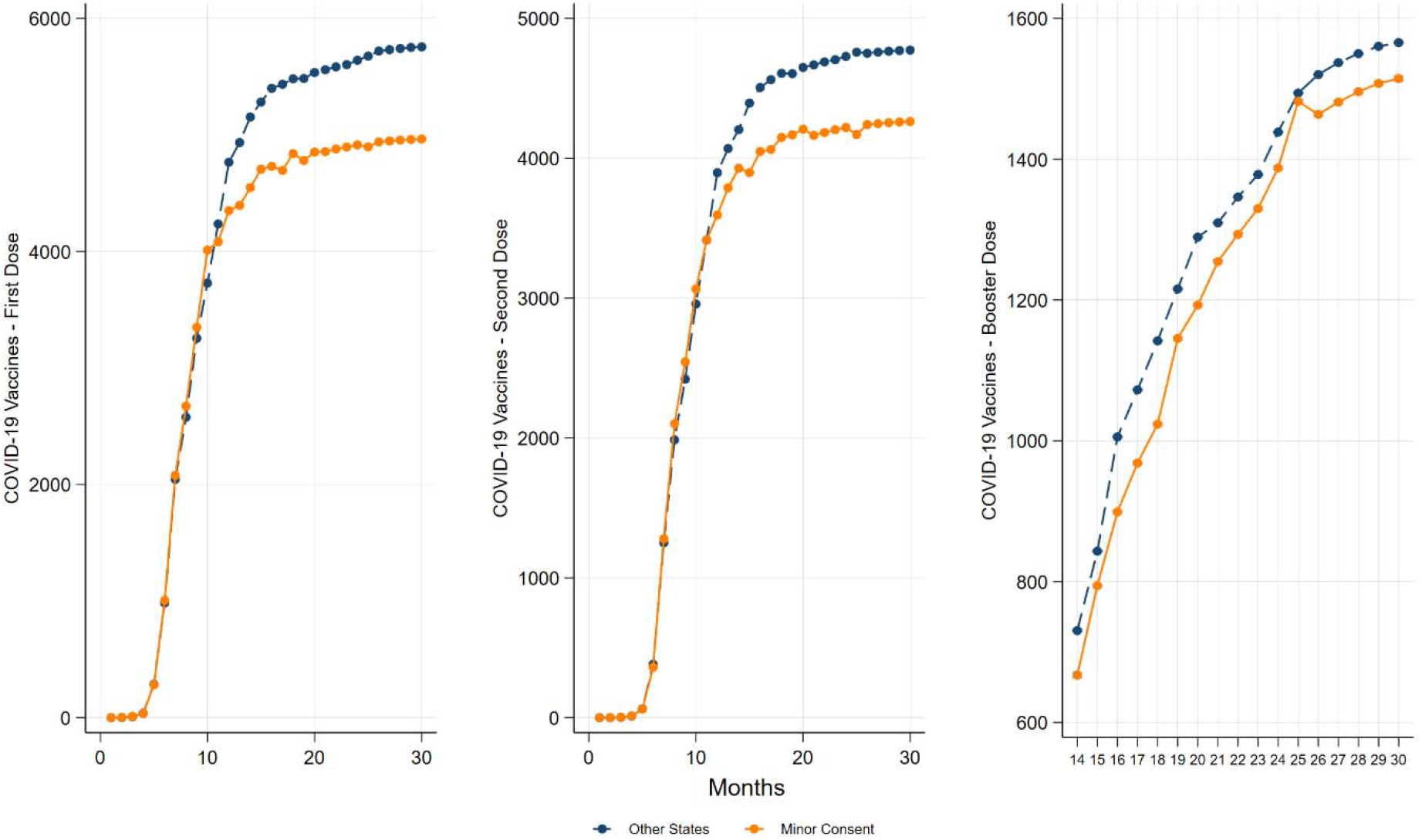
Vaccine Doses Administered Over Time. Figure 1 shows the population-adjusted number of COVID-19 vaccine doses administered to adolescents aged 12-17 based on data from the United States Centers for Disease Control and Prevention’s Immunization Information Systems. The x-axis represents weeks since December 2020. The y-axis represents doses administered.

**Figure 2.**
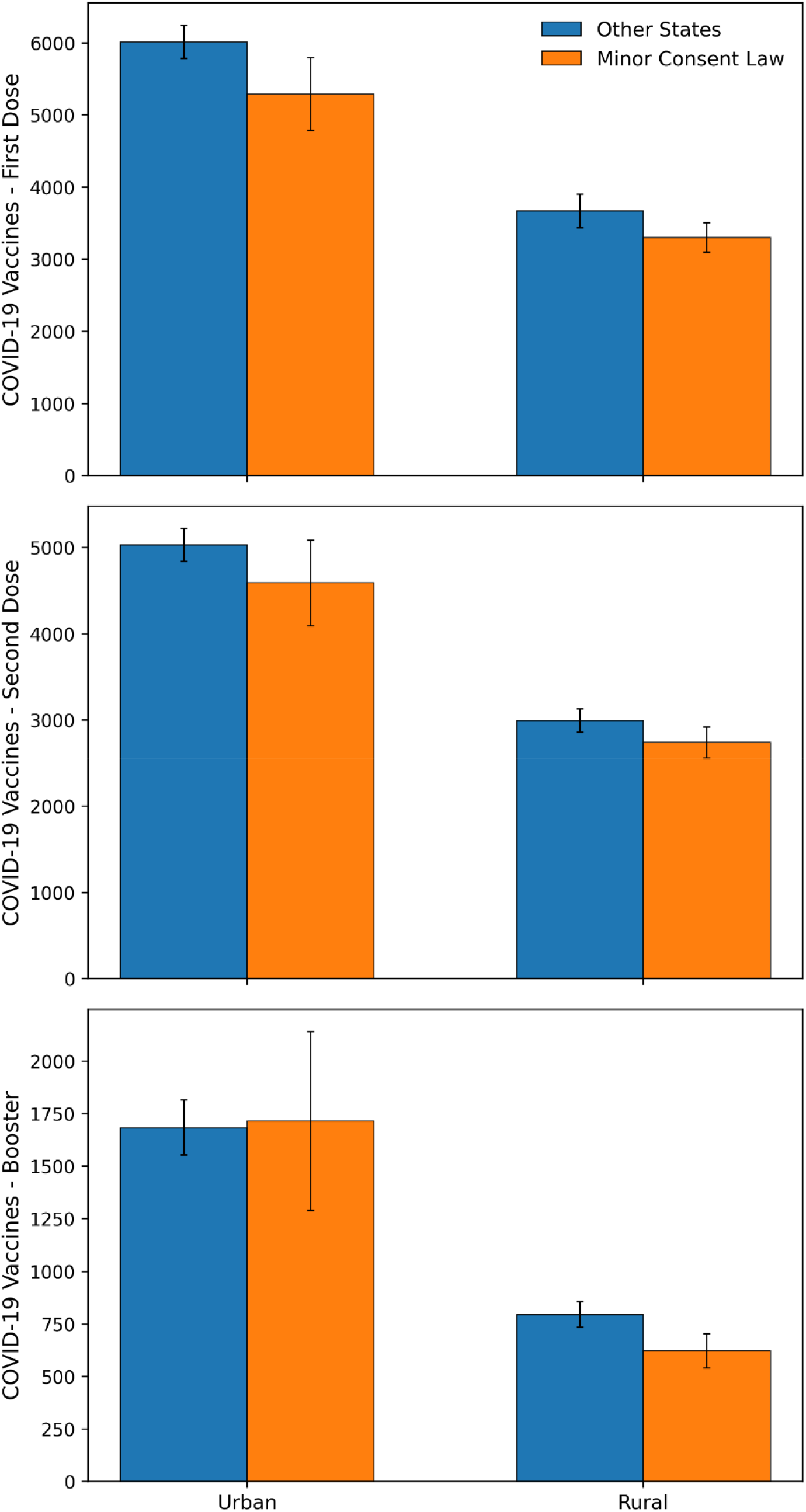
Vaccine Doses Administered in Urban vs. Rural Areas. Figure 2 shows the population-adjusted number of COVID-19 vaccine doses administered to adolescents aged 12-17 between December 2020 and May 2023 based on data from the United States Centers for Disease Control and Prevention’s Immunization Information Systems. Each vaccine dose type (initial, complete series, and booster) is separated by urban and rural areas and compares states with Minor Consent policies to states without.

### Poisson Regression Results

Overall, IRRs for the initial dose (IRR, 0.858; 95% CI: 0.718, 1.025; p = 0.0922) and complete series (IRR, 0.907; 95% CI: 0.706, 1.166; p = 0.4468) were not statistically significant. For booster doses, the IRR was 0.582 (95% CI: 0.409, 0.828; p = 0.0026). In urban counties, IRRs for the initial dose (IRR, 1.003; 95% CI: 0.743, 1.354; p = 0.9834), complete series (IRR, 0.960; 95% CI: 0.447, 2.066; p = 0.9176), and booster (IRR, 0.867; 95% CI: 0.722, 1.043; p = 0.1295) all were not statistically significant. In rural counties, IRRs for the initial dose (IRR, 0.871; 95% CI: 0.669, 1.134; p = 0.3063) and complete series (IRR, 0.993; 95% CI: 0.687, 1.435; p = 0.9705) were not statistically significant. However, in rural counties the IRR for booster doses was 0.541 (95% CI: 0.401, 730; p < 0.0001).

## Discussion

Minor Consent policies were not associated with increased adolescent COVID-19 vaccinations. Rather, Minor Consent policies were associated with lower booster doses in rural counties. This association is very much unlikely to be causal. This finding may relate to the changing attitudes towards adolescent vaccination over the course of the pandemic. The booster dose was authorized for ages sixteen and seventeen in December 2021, followed by ages twelve through fifteen in the following year^28^. Interviews with parents between 2022 and 2023 indicated that the number of parents who were “not at all hesitant” about the COVID vaccination for their child decreased, while the number who were “very hesitant” increased^9^. Broader shifts in parental opinion throughout the pandemic have also been documented, including decreased trust in the government, health care workers, and pharmaceutical companies^29^. Additionally, parents in communities with low COVID-19 vaccination rates were more likely to have their views of childhood vaccine effectiveness shift negatively during the pandemic^29^. How policies authorizing minors to consent to the COVID-19 booster related to these changing patterns could provide critical evidence on trust in vaccination programs beyond the pandemic.

It is unclear why Minor Consent policies were associated with lower booster uptake in rural but not urban areas. One possibility is differences in attitudes surrounding vaccination between rural and urban communities. Rural residents were more likely to oppose school and workplace vaccine mandates^30^. Minor Consent policies could be viewed similarly, and discussion of these policies potentially increase anti-vaccine sentiments. An example of backlash relating to Minor Consent policies has since been seen with changes to Tennessee’s Mature Minor Doctrine, which previously allowed adolescents to consent for COVID-19 vaccinations^23^. A memo put out after the vaccine was approved for children ultimately led to new legislation to clarify that minors cannot consent to immunizations^23^.

Minor Consent policies continue to be a topic of conversation for policymakers and public health advocates. Several states have implemented or introduced legislation allowing or revoking a minor’s authority to consent to vaccines. As these policies emerge, more research is needed to examine potential impacts on adolescent vaccine uptake. Additionally, how Minor Consent policies impact trust/hesitancy of parents of children or adolescents remains completely unknown. A greater understanding of these relationships could support the ongoing policy debate.

## Limitations

This study has several limitations. First, the analysis relied on an observational, non-experimental design with county-level administrative data, which cannot be used to draw causal inference. Second, Minor Consent policies were operationalized at the state level and may not capture heterogeneity in policy implementation, enforcement, or awareness, nor sub-state variation in access or consent practices. Third, the use of aggregated county-level data limits the ability to assess individual-level behaviors or mechanisms underlying vaccination patterns. Finally, there was no data to categorize whether minors consented for COVID-19 vaccines.

## Conclusions

Minor Consent policies for vaccinations have been around for decades. Unfortunately, most of the published studies have emphasized ethical and legal perspectives, leaving a void of empirical evidence evaluating these policies. Despite the lack of evidence, Minor Consent policies remain a topic of interest for state policymakers and public health advocates. Our current study revealed that Minor Consent policies for the COVID-19 vaccine were not associated with higher vaccination uptake in adolescents. Rather, for booster doses, we found that Minor Consent policies were associated with lower vaccination rates in rural counties. How the presence of Minor Consent policies relate to vaccination uptake, general vaccine hesitancy, and broader mistrust in public health institutions remains a critical area for future scientific inquiry. Understanding how Minor Consent policies influence parental trust in public health becomes an important step towards restoring the trust that has been eroding long before the pandemic began.

## Data Availability

COVID-19 vaccination data is publicly available at https://data.cdc.gov/Vaccinations/COVID-19-Vaccinations-in-the-United-States-County/8xkx-amqh/about_data. Analytic code can be found on publicly available repository [BLIND DURING PEER REVIEW].

## Ethics

Publicly available, deidentified secondary data does not meet definition of human subjects research. Conflicts: None to disclose.

## References

1. Johns Hopkins Coronavirus Resource Center. COVID-19 Overview. March 10, 2023. Accessed April 1, 2026. https://coronavirus.jhu.edu/region/united-states

2. Fitzpatrick M, Moghadas S, Pandey A, Galvani A. Two Years of U.S. COVID-19 Vaccines Have Prevented Millions of Hospitalizations and Deaths. The Commonwealth Fund; 2022. doi:10.26099/whsf-fp90

3. Wallace M. The Advisory Committee on Immunization Practices’ Interim Recommendation for Use of Pfizer-BioNTech COVID-19 Vaccine in Adolescents Aged 12–15 Years — United States, May 2021. MMWR Morb Mortal Wkly Rep. 2021;70. doi:10.15585/mmwr.mm7020e1

4. CDC. Archive: COVID-19 Vaccination and Case Trends by Age Group, United States | Data | Centers for Disease Control and Prevention. Accessed April 1, 2026. https://data.cdc.gov/Vaccinations/Archive-COVID-19-Vaccination-and-Case-Trends-by-Ag/gxj9-t96f/data_preview

5. CDC. COVID-19 Vaccination Coverage and Intent for Vaccination, Children 6 months through 17 years, United States. Published online December 2, 2025. Accessed April 1, 2026. https://www.cdc.gov/covidvaxview/weekly-dashboard/child-coverage-vaccination.html

6. Saelee R. Disparities in COVID-19 Vaccination Coverage Between Urban and Rural Counties — United States, December 14, 2020–January 31, 2022. MMWR Morb Mortal Wkly Rep. 2022;71. doi:10.15585/mmwr.mm7109a2

7. Ruiz JB, Bell RA. Parental COVID-19 Vaccine Hesitancy in the United States. Public Health Rep. 2022;137(6):1162–1169. doi:10.1177/00333549221114346

8. Nguyen KH, Nguyen K, Mansfield K, Allen JD, Corlin L. Child and adolescent COVID-19 vaccination status and reasons for non-vaccination by parental vaccination status. Public Health. 2022;209:82–89. doi:10.1016/j.puhe.2022.06.002

9. Santibanez TA, Black CL, Zhou T, Srivastav A, Singleton JA. Parental hesitancy about COVID-19, influenza, HPV, and other childhood vaccines. Vaccine. 2024;42(25):126139. doi:10.1016/j.vaccine.2024.07.040

10. Mihaly LK, Schapiro NA, English A. From Human Papillomavirus to COVID-19: Adolescent Autonomy and Minor Consent for Vaccines. Journal of Pediatric Health Care. 2022;36(6):607–610. doi:10.1016/j.pedhc.2022.06.007

11. Sharko M, Jameson R, Ancker JS, Krams L, Webber EC, Rosenbloom ST. State-by-State Variability in Adolescent Privacy Laws. Pediatrics. 2022;149(6):e2021053458. doi:10.1542/peds.2021-053458

12. Olick RS, Yang YT, Shaw J. Adolescent Consent to COVID-19 Vaccination: The Need for Law Reform. Public Health Rep. 2021;137(1):163–167. doi:10.1177/00333549211048784

13. Sawyer K, Rosenberg AR. How Should Adolescent Health Decision-Making Authority Be Shared? AMA Journal of Ethics. 2020;22(5):372–379. doi:10.1001/amajethics.2020.372

14. Morgan L, Schwartz JL, Sisti DA. COVID-19 Vaccination of Minors Without Parental Consent: Respecting Emerging Autonomy and Advancing Public Health. JAMA Pediatr. 2021;175(10):995–996. doi:10.1001/jamapediatrics.2021.1855

15. Moore CM, Wakim PG, Taylor HA. Factors Affecting COVID-19 Vaccine Decision-Making and Satisfaction: A Survey of U.S. High School Students. Journal of Adolescent Health. 2024;74(6):1139–1145. doi:10.1016/j.jadohealth.2024.01.025

16. Rogers AA, Cook RE, Button JA. Parent and Peer Norms are Unique Correlates of COVID-19 Vaccine Intentions in a Diverse Sample of U.S. Adolescents. Journal of Adolescent Health. 2021;69(6):910–916. doi:10.1016/j.jadohealth.2021.09.012

17. Torres AR, Johnson NP, Ellingson MK, et al. State Laws Permitting Adolescent Consent to Human Papillomavirus Vaccination and Rates of Immunization. JAMA Pediatr. 2022;176(2):203–205. doi:10.1001/jamapediatrics.2021.4591

18. Silverman RD, Opel DJ, Omer SB. Vaccination over Parental Objection — Should Adolescents Be Allowed to Consent to Receiving Vaccines? New England Journal of Medicine. 2019;381(2):104–106. doi:10.1056/NEJMp1905814

19. Opsahl R. House subcommittee approves bill requiring parental consent for HPV vaccine . Iowa Capital Dispatch. Iowa Capital Dispatch. January 14, 2026. Accessed April 1, 2026. https://iowacapitaldispatch.com/2026/01/14/house-subcommittee-approves-bill-requiring-parental-consent-for-hpv-vaccine/

20. Fisher H, Harding S, Hickman M, Macleod J, Audrey S. Barriers and enablers to adolescent self-consent for vaccination: A mixed-methods evidence synthesis. Vaccine. 2019;37(3):417–429. doi:10.1016/j.vaccine.2018.12.007

21. Irwin M, Soled DR, Cummings CL. Lowering the Age of Consent for Vaccination to Promote Pediatric Vaccination: It’s Worth a Shot. J Law Med Ethics. 2024;52(1):52–61. doi:10.1017/jme.2024.43

22. Fact Sheet: Colorado Minors Can Now Access Some Vaccines on Their Own-How A New Law Makes This Possible-Immunize Colorado. https://www.immunizecolorado.org/. Accessed April 1, 2026. https://www.immunizecolorado.org/resources/fact-sheet-colorado-minors-can-now-access-some-vaccines-on-their-own-how-a-new-law-makes-this-possible/

23. Hijano DR, Yaun JA. Mature Minor Doctrine Clarification Act: A Setback in Pediatric Immunizations. J Pediatric Infect Dis Soc. 2024;13(2):155–158. doi:10.1093/jpids/piae006

24. CDC. COVID-19 Vaccinations in the United States,County. Published online May 12, 2023. Accessed April 1, 2026. https://data.cdc.gov/Vaccinations/COVID-19-Vaccinations-in-the-United-States-County/8xkx-amqh/about_data

25. ERS. Rural-Urban Continuum Codes. Accessed April 1, 2026. https://www.ers.usda.gov/data-products/rural-urban-continuum-codes

26. Teens for Vaccines. Minor Consent Laws for Vaccinations by State. teens for vaccines. 2026. Accessed April 1, 2026. https://teensforvaccines.org/minor-consent-laws-by-state/

27. VaxTeen. Consent Laws by State — VaxTeen. July 2025. Accessed April 1, 2026. https://web.archive.org/web/20250709220625/ https://www.vaxteen.org/consent-laws-by-state

28. FDA. Coronavirus (COVID-19) Update: FDA Expands Eligibility for Pfizer-BioNTech COVID-19 Booster Dose to 16-and 17-Year-Olds. FDA. August 9, 2024. Accessed April 1, 2026. https://www.fda.gov/news-events/press-announcements/coronavirus-covid-19-update-fda-expands-eligibility-pfizer-biontech-covid-19-booster-dose-16-and-17

29. Grills LA, Wagner AL. The impact of the COVID-19 pandemic on parental vaccine hesitancy: A cross-sectional survey. Vaccine. 2023;41(41):6127–6133. doi:10.1016/j.vaccine.2023.08.044

30. Sparks G, Hamel L, Kirzinger A, Stokes M, Brodie M. KFF COVID-19 Vaccine Monitor: Differences in Vaccine Attitudes Between Rural, Suburban, and Urban Areas. KFF; 2021. Accessed April 1, 2026. https://www.kff.org/covid-19/kff-covid-19-vaccine-monitor-vaccine-attitudes-rural-suburban-urban/

